# Cervical Whole Slide Histology Image Analysis Toolbox

**DOI:** 10.1101/2020.07.22.20160366

**Authors:** Sudhir Sornapudi, Ravitej Addanki, R. Joe Stanley, William V. Stoecker, Rodney Long, Rosemary Zuna, Shellaine R. Frazier, Sameer Antani

## Abstract

Cervical intraepithelial neoplasia (CIN) is regarded as a potential precancerous state of the uterine cervix. Timely and appropriate early treatment of CIN can help reduce cervical cancer mortality. Accurate estimation of CIN grade correlated with human papillomavirus (HPV) type, which is the primary cause of the disease, helps determine the patient’s risk for developing the disease. Colposcopy is used to select women for biopsy. Expert pathologists examine the biopsied cervical epithelial tissue under a microscope. The examination can take a long time and is prone to error and often results in high inter- and intra-observer variability in outcomes. We propose a novel image analysis toolbox that can automate CIN diagnosis using whole slide image (digitized biopsies) of cervical tissue samples. The toolbox is built as a four-step deep learning model that detects the epithelium regions, segments the detected epithelial portions, analyzes local vertical segment regions, and finally classifies each epithelium block with localized attention. We propose an epithelium detection network in this study and make use of our earlier research on epithelium segmentation and CIN classification to complete the design of the end-to-end CIN diagnosis toolbox. The results show that automated epithelium detection and segmentation for CIN classification yields comparable results to manually segmented epithelium CIN classification. This highlights the potential as a tool for automated digitized histology slide image analysis to assist expert pathologists.

## I. INTRODUCTION

Cervical cancer is widely occurring cancer and a major health problem in women worldwide. It is usually caused by sexually transmitted infections from certain types of Human Papillomavirus (HPV). According to WHO [1], in 2018, cervical cancer was recorded as the second most common cancer in women in low and middle-income regions with an estimated 570,000 new cases and approximately 311,000 deaths occurring during that year [2]. Women aged 20 to 39 years are more vulnerable accounting for 10 premature deaths per week [3]. However, early stage diagnosis can help prevent cervical cancer.

Tissue specimens from the uterine cervix of affected women is extracted through biopsy and affixed on glass slides and stained with hematoxylin and eosin (H&E). Then, an expert histopathologist examines the glass sides under a light microscope to provide the diagnosis for each sample, as shown in Figure 1. Accurate interpretation of glass slides is crucial to avoid misdiagnoses [4], which requires extensive time and effort by the pathologist. Each woman could have up to a dozen biopsy samples that require analysis. This displays the necessity of computational digital pathology to augment and automate the process of diagnosis by scanning the digitalized whole slide image (WSI) [5][6].

**Figure 1.**
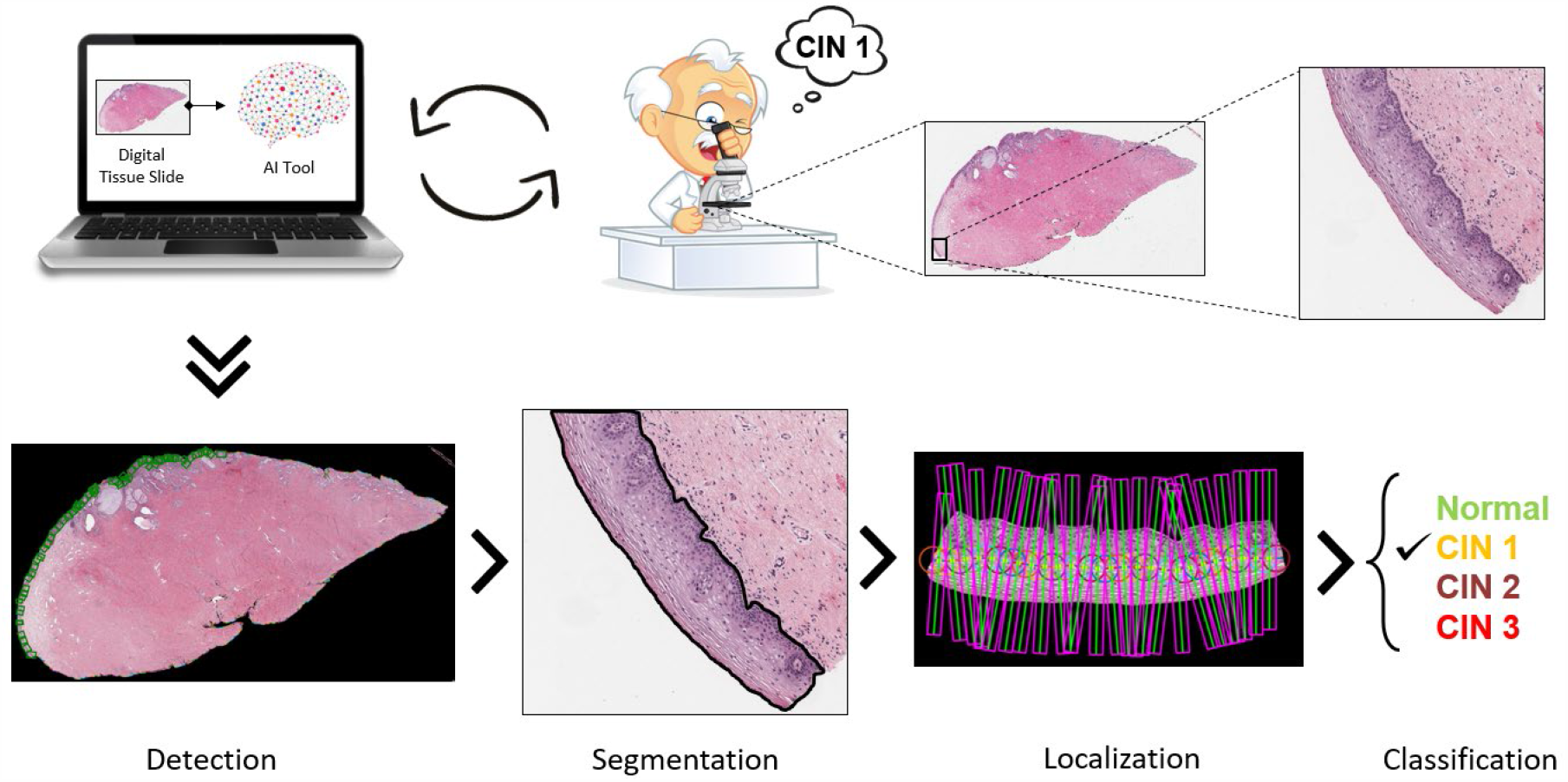
Graphical overview of the proposed toolbox.

Grading of cervical disease is largely based on the proportion of immature cells within the epithelium, starting at the base and gradually encompassing the entire epithelial layer. This pre-cancerous condition is called Cervical Intraepithelial Neoplasia (CIN) or cervical dysplasia. The CIN lesions are caused by HPV. All the cells in the epithelium contain the virus, but the degree to which the epithelium can mature is dependent on the degree to which the virus interferes with the cellular maturation process. The disease is present in the entire epithelial layer in all cases, and the degree of differentiation determines the grade. In CIN1, the immature cells are in the lower one-third; in CIN2, they are found in the lower two-third, and in CIN3, they are found in the entire layer. But the cells at the top are different. They are larger in CIN1 because the tissue can mature to a higher level than the other grades. In CIN3, there is little or no maturation, so the cells at the top look very similar to those at the base [7].

The histopathological WSIs have some unique challenges to overcome. The sheer size of WSI data contains billions of pixels, comprising gigabytes of data. There is a high variability of image appearance due to slide preparation, staining, and various other artefacts during the scanning of the tissue slides. Additionally, the shapes of the biopsied tissue samples vary, and there is no standard shape and size of the epithelium regions and the abnormal cells present inside these regions. The presence of blood stains, ink markers, tapes, and blurred regions pose challenges when designing automated tools. These problems present unique barriers to the development of deep learning models in digital pathology. Nonetheless, the use of deep learning (DL) methods in digital pathology has been proven to have a significant improvement in diagnostic capabilities and efficiency [8][9][10]. The histopathological analysis is performed for various diseases like cervical cancer, skin cancer, breast cancer, prostate cancer, etc. The effects of stain color augmentation and stain color normalization are studied, and an unsupervised approach for stain color normalization was proposed using neural networks for computational pathology [11]. The use of convolution neural networks (CNN) for segmentation, detection, and classification in common histologic primitives were explored by Janowczyk *et al*. [12]. Multi-instance learning is proposed for image-level classification and annotating relevant regions for histology image analysis [13]. Focusing on cervical cancer, Wang *et al*. [14] presented a block segmentation method to extract textural feature information for CIN classification using support vector machines. Superpixel-based DL nuclei detection was explored in cervical histology images [15]. The problems of inter-observer variability and the advantages of the use of computer-aided systems as a secondary decision for classifying precursor lesions were presented by Albayrak *et al*. [16]. Li *et al*. detailed the use of various machine learning techniques for cervical histopathology image analysis.

Our current study leverages various deep learning models and specifically seeks to automate the diagnosis of cervical cancer by scanning histopathological WSIs. This is an end-to-end prototype tool that assists pathologists with valuable information like the location of epithelium regions and can also coarsely segment the epithelium regions from the background and unwanted tissue regions and classify these epithelium regions with added contributions of local regions for the overall classification. We introduce epithelium detection with this study and utilize our previous work on epithelium segmentation [17] and CIN classification [18] to design the toolbox.

This is a novel toolbox that is inspired from the way pathologists analyze the glass slides under a microscope: looking along the outer edges of the tissue and identifying the epithelium regions; zooming in and observing the cell distribution and patterns across the epithelium in detail; and quantifying the CIN grades along the epithelium regions as depicted in Figure 1 and Figure 2.

**Figure 2.**
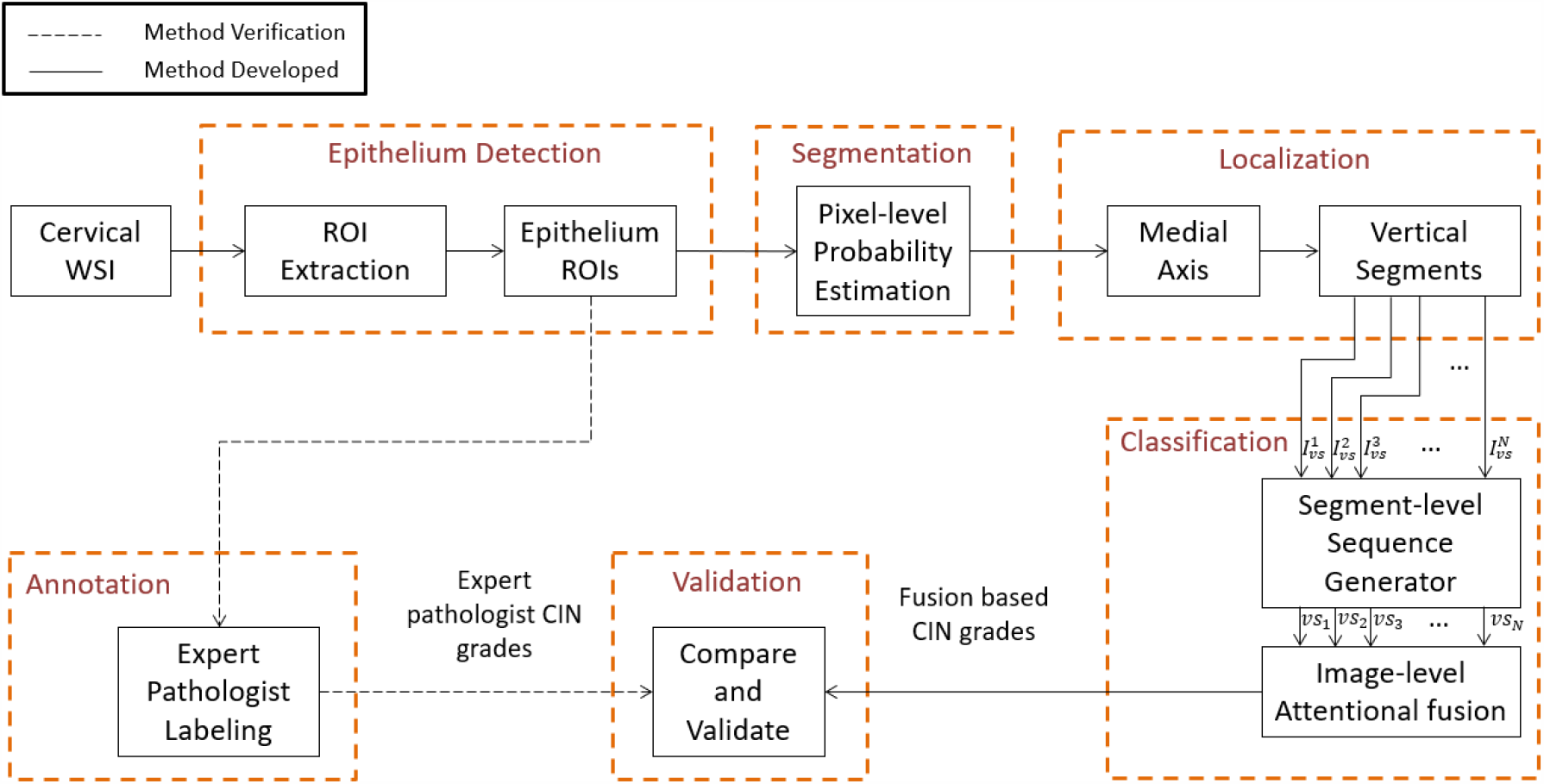
Overview of the proposed toolbox

## II. METHODOLOGY

The toolbox incorporates a four-step sequential procedure (Figure 2). First, the outer region of interest (ROI) is identified, and the regions are filtered with the epithelium detection network. Second, pixel-level epithelium segmentation takes place. Third, localization occurs to generate vertical segments. Fourth, CIN grade classification with attention-based sequential feature modeling is completed.

### Epithelium Detection

We propose the epithelium detection process with an initial preprocessing that includes the extraction of ROIs from the low-resolution WSIs (refer Section 3.1). This is followed by a classifier network that identifies the epithelium ROIs by reading the high-resolution versions (refer Section 3.1) of the extracted ROIs.

### ROI Extraction

Initially, we process the low-resolution version of raw cervical histology WSI to generate a mask for the tissue region, determine the contour, and draw boxes around the outer region of the digitized tissue sample. The WSIs usually have a tissue specimen with a white background. Since the background is uniform, a simple threshold operation can create a mask for the WSI. This mask is further processed to remove small unwanted object regions and close the holes in the object regions.

Instead of using grid-based region creation, we optimize the selection of epithelium regions by focusing only on the outer regions, where the epithelium layer is present. The contour of the mask provides the outer edge information. This contour curve is cut into a piece-wise curve at a frequency of 40 points per cut (chosen empirically based on the low-resolution slide images). In order to draw boxes of ROIs, a polygon is fit based on the points from each piece-wise curve and a tangent is drawn at the midpoint of these piecewise curves. Based on the tangential lines, rectangular boxes were drawn facing the object region of the mask, as shown in Figure 3. The width of the ROI is determined by the maximum and the minimum values of horizontal coordinates and the height is chosen to be 40 pixels (chosen empirically) to accommodate the entire epithelium cross-section. These rectangular box coordinates were normalized and recorded. The high-resolution ROIs (at 10× magnification) were finally created by cropping out the image regions from the high-resolution slide image using the normalized rectangular bounding box coordinates data as shown in Figure 4.

**Figure 3.**
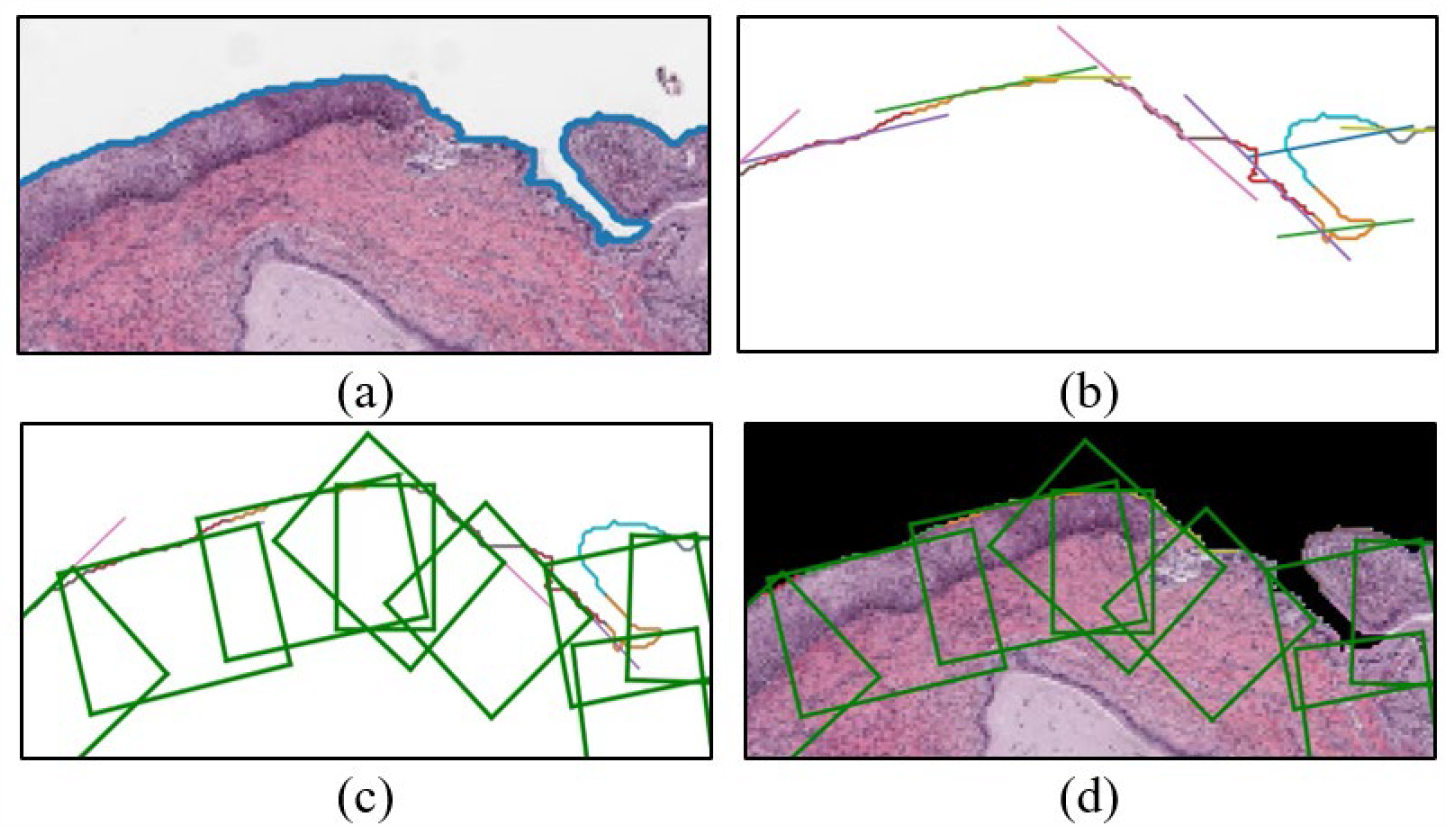
Steps for ROI extraction. (a) Finding the contour on the edge of the tissue sample, (b) Piece-wise curve for drawing tangents, (c) rectangular boxes drawn with reference to tangents, and (d) ROI boxes on the original masked image.

**Figure 4.**
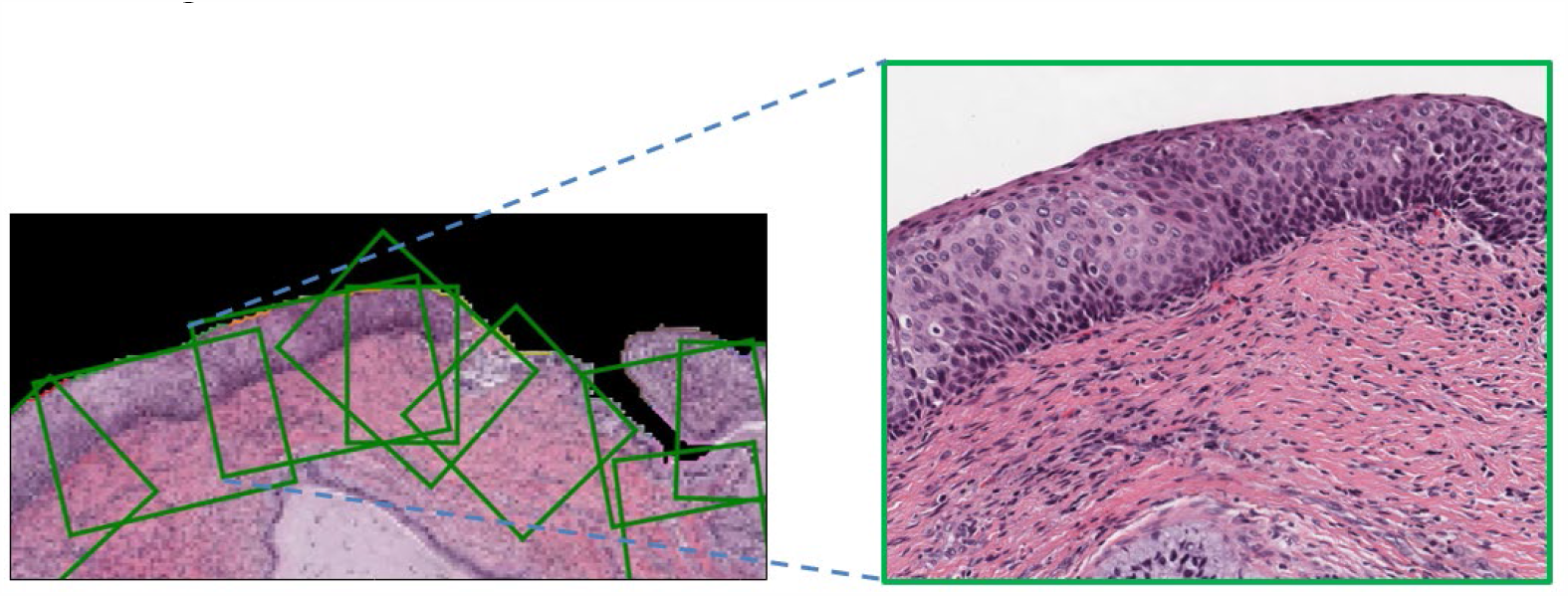
Mapping of high-resolution ROI (right) to its low-resolution image (left).

### Epithelium Detection Network

The epithelium detection network is a binary classifier that categories an input image as epithelium or non-epithelium. The high-resolution ROIs are fed to this network to filter and retain only the epithelium containing ROIs, as shown in Figure 5.

**Figure 5.**
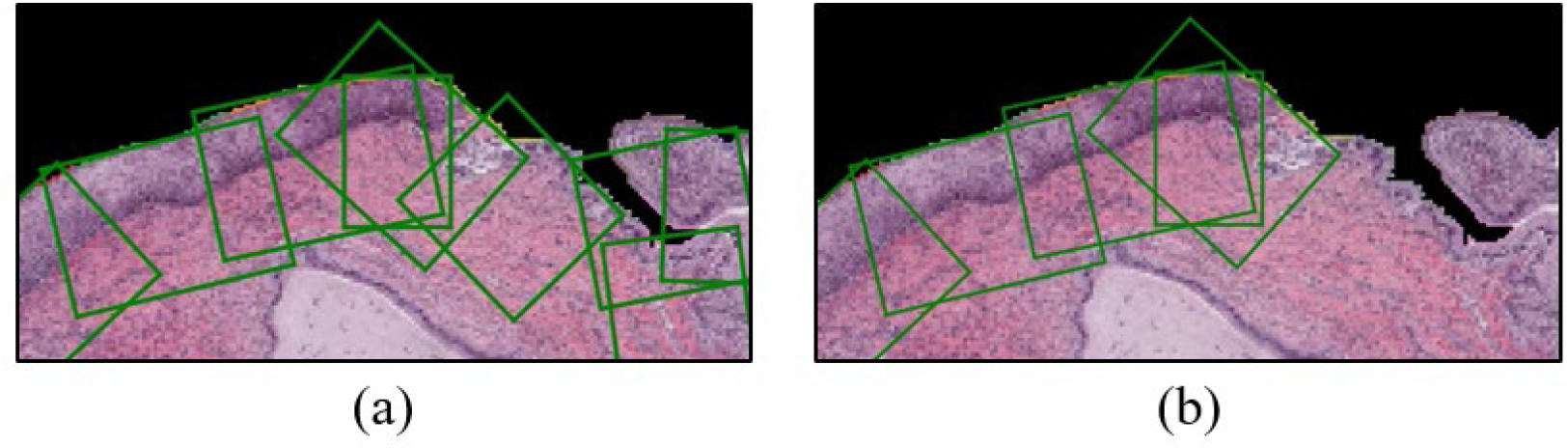
Filtering of epithelium ROIs with the results from the epithelium detection network.

Table 1 presents the network architecture that was investigated. This is a customized version of the VGG-19 model [19], where the initial layers contain a series of convolution block and max-pooling layers. The end feature maps generated from these layers are vectorized and passed through fully connected layers. All the aforementioned layers were activated with ReLU non-linearity functions, except the last fully connected layer that contains two neurons to compute the classification probability for each class using the SoftMax activation function. To reduce overfitting, the output of the first fully connected layer is constrained by randomly dropping 50% of the values to zero.

**Table 1.** Epithelium Detection Network Architecture

| Layers | Configurations | Size |
| --- | --- | --- |
| Input | - | $3 \times 250 \times 250$ |
| Conv Block 1 | $[k: 3 \times 3, s: 1, p: 1] \times 2$ | $64 \times 250 \times 250$ |
| Pool 1 | $mp: 2 \times 2, s: 2$ | $64 \times 125 \times 125$ |
| Conv Block 2 | $[k: 3 \times 3, s: 1, p: 1] \times 2$ | $128 \times 125 \times 125$ |
| Pool 2 | $mp: 2 \times 2, s: 2$ | $128 \times 62 \times 62$ |
| Conv Block 3 | $[k: 3 \times 3, s: 1, p: 1] \times 4$ | $256 \times 62 \times 62$ |
| Pool 3 | $mp: 2 \times 2, s: 2$ | $256 \times 31 \times 31$ |
| Conv Block 4 | $[k: 3 \times 3, s: 1, p: 1] \times 4$ | $512 \times 31 \times 31$ |
| Pool 4 | $mp: 2 \times 2, s: 2$ | $512 \times 15 \times 15$ |
| Conv Block 5 | $[k: 3 \times 3, s: 1, p: 1] \times 4$ | $512 \times 15 \times 15$ |
| Pool 5 | $mp: 2 \times 2, s: 2$ | $512 \times 7 \times 7$ |
| Flatten | - | $25088 \times 1$ |
| FC 1 | $nh: 1024$ | $1024 \times 1$ |
| Dropout | $prob: 0.5$ | $1024 \times 1$ |
| FC 2 | $nh: 1024$ | $1024 \times 1$ |
| FC 3 | $nh: 2$ | $2 \times 1$ |
| Output | $softmax$ | $2 \times 1$ |

$k, s, p, mp, nh, prob$ are kernel, stride size, padding size, max pooling, number of neurons, and probability, respectively. ‘FC’ denotes a fully connected single-layer neural network.

The weights in the convolutional layers are initialized with Kaiming initialization [20] for better stability, and the fully connected layers are initialized with the normal distribution. In the training phase, the weights are iteratively updated with the gradients of the cross-entropy loss function, which is computed via RMSprop optimization over a mini-batch of training samples. The initial learning rate is set to 0.0001 and changes adaptively as the training progresses.

### Epithelium Segmentation

From Figure 2, epithelium segmentation is the second step in the slide analysis process. We utilize the EpithNet model from our previous studies [17] to coarsely segment the high-resolution epithelium ROIs to generate an epithelium segmentation mask. The segmentation model is a pixel-wise epithelial probability estimator and is developed based on the information provided by a pixel depending on the surrounding spatial proximity in the image plane. The epithelium ROI is preprocessed by splitting into tiles, and each tile is further processed to generate 64×64×3 RGB patch image data. These patches are created with a sliding window technique with stride 4. From [17], the EpithNet-64 regression model was utilized to process these patch data to output an estimated probability of the center pixel of being an epithelium. These pixel probabilities are gathered and treated as pixel intensities to finally form a mask. This mask is post-processed by applying thresholding, morphology, and smoothing filters to finally generate a binary segmentation mask.

### Localization

CIN is the growth of atypical cervical cells in the epithelium. This abnormal growth is clearly understood when observed locally. Thus, standard width vertical segments [18] are generated from the epithelium ROIs with reference to the medial axis, drawn with the help of epithelium segmentation mask information, as shown in Figure 6. The details are provided in our previous work [18].

**Figure 6.**
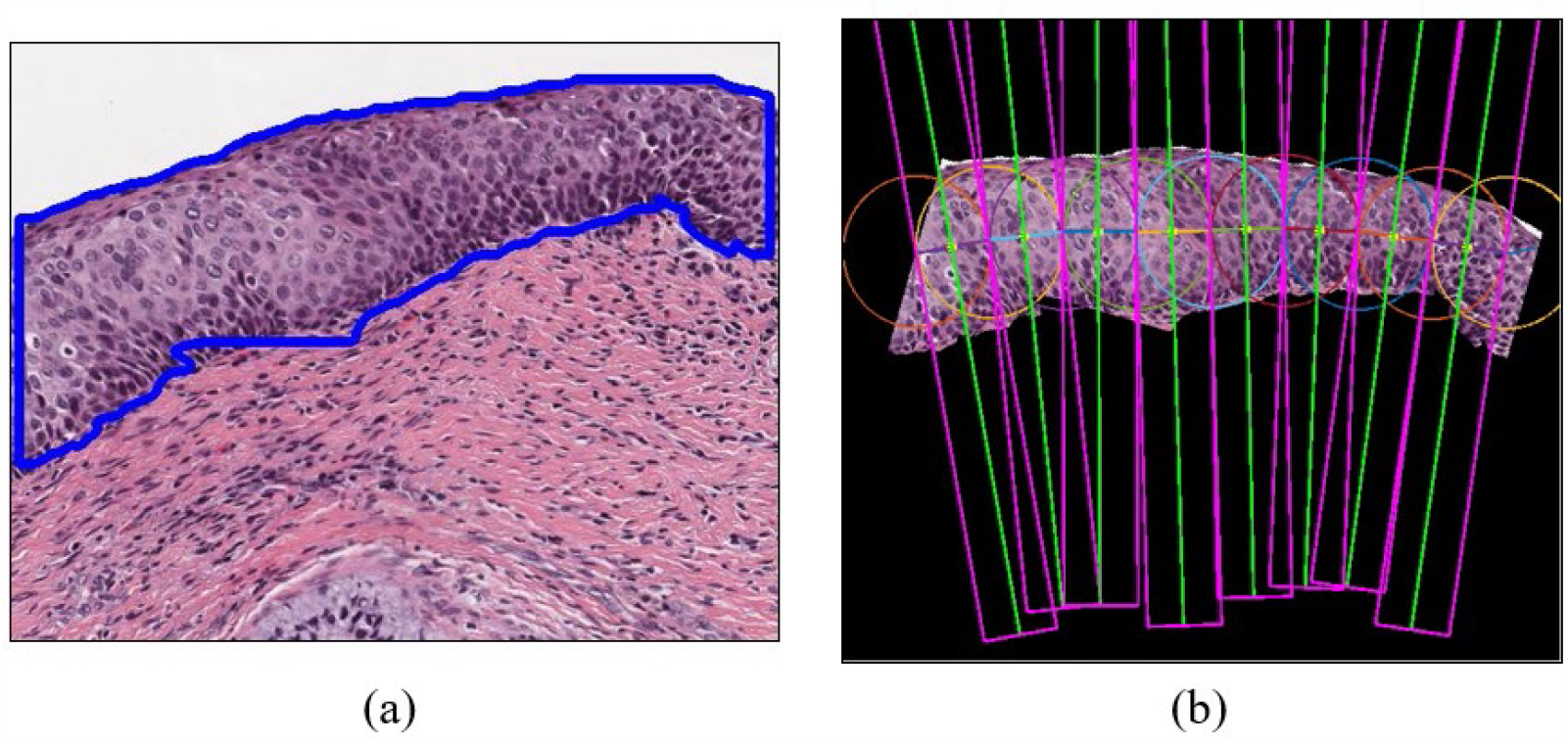
(a) Epithelium segmentation mask overlaid as a contour on the epithelium ROI. (b) Vertical segments generation through the localization process.

### CIN Classification

For each segmented epithelium, CIN classification is performed. The DeepCIN [18] is a two-fold learning process. First, a segment-level sequence generator is a weakly supervised network that scans each localized vertical segment image to generate the best sequence representation of the input image. This is built as an encoder-decoder model, where the encoder is a CNN model that extracts and encodes convolutional spatial feature information to a sequential feature. The decoder is a many-to-one model that consists of two layers of bidirectional Long-Short-Term Memory (BLSTM) network and a single layer neural network. Second, an attention-based fusion network is an image-level classifier that sequentially interprets the vertical segment sequences. This provides a contextual understanding of local information that not only helps in providing the multi-class CIN classification result, but also provides the contribution of each vertical segment towards the final classification. This is built with gated recurrent units (GRUs) and attentional neural network layers. The detailed model implementation can be found in the work of Sornapudi *et al*. [18].

The model was previously trained with 5-fold cross-validation, and we ensemble the five trained models to produce the CIN classification result on the proposed toolbox.

## III. EXPERIMENTS

### Data

The study uses 150 H&E stained cervical histopathological slides (WSI) provided by the Department of Pathology at the University of Oklahoma Medical Center in collaboration with the National Library of Medicine. The glass slides were scanned using the Aperio ScanScope slide scanner with 20× objective, producing WSIs in a pyramidal tiled format with the file extension “svs”. These SVS files are large (they typically range from 1 gigabyte to 100 megabytes). Each pixel has a size of 0.25 µm^2^. The pyramidal tile level varies from 0 to 2/3/4. In this paper, we often refer to a 1× magnification image (highest pyramid level) as a low-resolution image and 20× magnification image (pyramid level 0) down sampled to 10× magnification as a high-resolution image. This is explicitly performed to maintain the same image resolutions used in our previous works [17][18]. There are three sets of WSIs captured during the years 2013, 2015, and 2016, and hence named OU13, OU15, and OU16, respectively. Each of these sets contains 50 WSIs. The study uses 50 WSIs from the OU13 set for training and validation of the epithelium detection model, and 100 WSIs from both the OU15 set and the OU16 set for testing our toolbox. With our automated ROI extraction technique, we could generate high-resolution arbitrary size images that contain epithelium and non-epithelium regions. The distribution of the image blocks can be observed in Table 2. The images from Table 2 will be evaluated for the correctness of the epithelium segmentation process in Section 4.1.

**Table 2.** Data Distribution for Epithelium detection

| Dataset | WSIs | Epithelium ROIs | Non-epithelium ROIs |
| --- | --- | --- | --- |
| OU13 | 50 | 2,998 | 20,841 |
| OU15 | 50 | 4,915 | 12,595 |
| OU16 | 50 | 4,106 | 8,601 |

The data set examined in this research for evaluating CIN classification model (Figure 2) consists of a total of 947 expert-labeled epithelium images (a subset of obtained epithelium ROIs), 723 images from OU15-set, and 224 images from OU16-set. The class distribution of the data is shown in Table 3. It should be noted that these 947 epithelium images are an independent set of images extracted from the proposed approach and are mutually exclusive from the manually extracted epithelium images that are used for training the CIN classification model [18].

**Table 3.** Subset of epithelium ROI images for evaluating CIN classification

| Class | OU15 | OU16 | Combined set |
| --- | --- | --- | --- |
| Normal | 451 | 133 | 584 |
| CIN 1 | 90 | 11 | 101 |
| CIN 2 | 128 | 41 | 169 |
| CIN 3 | 54 | 39 | 93 |
| Total | 723 | 224 | 947 |

### Implementation Details

The architecture of the epithelium detection network is summarized in Table 1. The network incorporates a transfer learning scheme. So, the entire model is pre-trained on the ImageNet classification dataset. The convolutional module weights were frozen, and the rest of the layer weights were reinitialized with random Gaussian distributions.

We have designed the CNN model such that it can read RGB input images of size 250×250×3. To maintain a standard resolution of the input images, the extracted ROIs are padded with zeros, center cropped to size 500×500×3 and finally resized to 250×250×3. We incorporated data augmentation techniques to avoid the problem of highly imbalanced data in the training set (Table 2). The epithelium ROI images are upsampled to count equivalent to non-epithelium ROIs (20,841) via augmentations like random rotate, vertical and horizontal flip, random blur, etc. RMSprop, with a mini-batch of size 32, is used to train the network for 100 epochs. Early stopping is applied to monitor the generalization error and avoid overfitting.

In the testing phase, the ROIs categorized as epithelium were further processed with previously trained models in the toolbox: EpithNet-64 and DeepCIN. We obtain an epithelium segmentation mask with EpithNet-64 for the generation of vertical segments that are consumed by DeepCIN to deliver the CIN classification results and the contribution of the vertical segments towards the classification output. The models are run on the PyTorch v1.4 platform [21] using Nvidia Quadro P4000 GPU with 8GB of memory.

### Evaluation Metrics

We evaluate the proposed epithelium detection network for classification as epithelium/non-epithelium ROIs. The performance evaluation metrics include specificity (*Sp*), sensitivity (*Se*), harmonic mean (*H*_*mean*_), F1-score (*ACC*), accuracy (*AUC*), and area under the ROC curve (*AUC*). *Sp* measures the proportion of correctly identified non-epithelium ROIs, *Se* measures the proportion of correctly identified epithelium ROIs, *H*_*mean*_ is the harmonic mean of *Sp* and *Se* (which is better at measuring under imbalance data distribution), *F*1 is the harmonic mean of precision and recall, and *ACC* is the global accuracy. *AUC* is the area under the receiver operating characteristic curve and is plotted with varying thresholds on final classification scores.

We also evaluate the final CIN classification results from the detected epithelium ROIs. The scoring metrics used are precision (*P*), recall (*R*), F1-score (*F*1), classification accuracy (*ACC*), area under Receiver Operating Characteristic curve (*AUC*), average precision (*AP*), Matthews correlation coefficient (*MCC*), and Cohen’s kappa score (κ) [18]. The percentage weighted average scores were computed to account for the imbalance in the data distribution.

## IV. RESULTS

We evaluate the toolbox performance by comparing the epithelium detection network results and the CIN classification results against the expert pathologist annotated ground truths on the OU15 and OU16 WSI datasets.

### Performance of Epithelium Detection Network

Table 4 shows the classification performance (*Sp, Se, H*_*mean*_, *FF*1, *ACC*, and *AUC*) of the proposed epithelium detection network. The objective of this network is to sort the extracted ROI images into epithelium and non-epithelium. Since there are more non-epithelium ROIs compared to epithelium ROIs (Table II), the specificity is always observed to be higher than sensitivity. Harmonic mean (*H*_*mean*_) gives a better-balanced score between *Sp* and *Se*, and is found to have a mean value of 97.3%, 92.7%, and 95.0% among OU15, OU16, and OU15 and OU16 combined datasets, respectively. We observed that the trained network has better generalization on the OU15-set, compared to the OU16-set. The combined dataset results were also reported. We could not compare the performance of the network with other works because, to our knowledge, this is the first study on cervical epithelium detection.

**Table 4.** Epithelium Detection Results

| Test set | $Sp$ | $Se$ | $H_{mean}$ | $F1$ | $ACC$ | $AUC$ |
| --- | --- | --- | --- | --- | --- | --- |
| OU15 | 98.3 | 96.6 | 97.3 | 95.6 | 97.8 | 97.4 |
| OU16 | 96.3 | 90.8 | 92.7 | 91.4 | 95.2 | 93.5 |
| OU15/OU16 | 97.3 | 93.7 | 95.0 | 93.5 | 96.5 | 95.5 |

Figure 7 contains examples of correctly classified epithelium ROIs (true positive) and misclassified epithelium ROIs (false positive). Typically, the cancer cells are manifested in the epithelium, and hence the identification of epithelium is our top priority. The network is observed to identify the epithelium regions even under challenging conditions. The falsely identified ROIs closely resemble the epithelium regions, which makes the classification task difficult. Nevertheless, the network has provided good performance accuracy results of 97.8% on OU15-set, 95.2% on OU-16, and 96.5% on the combined set.

**Figure 7.**
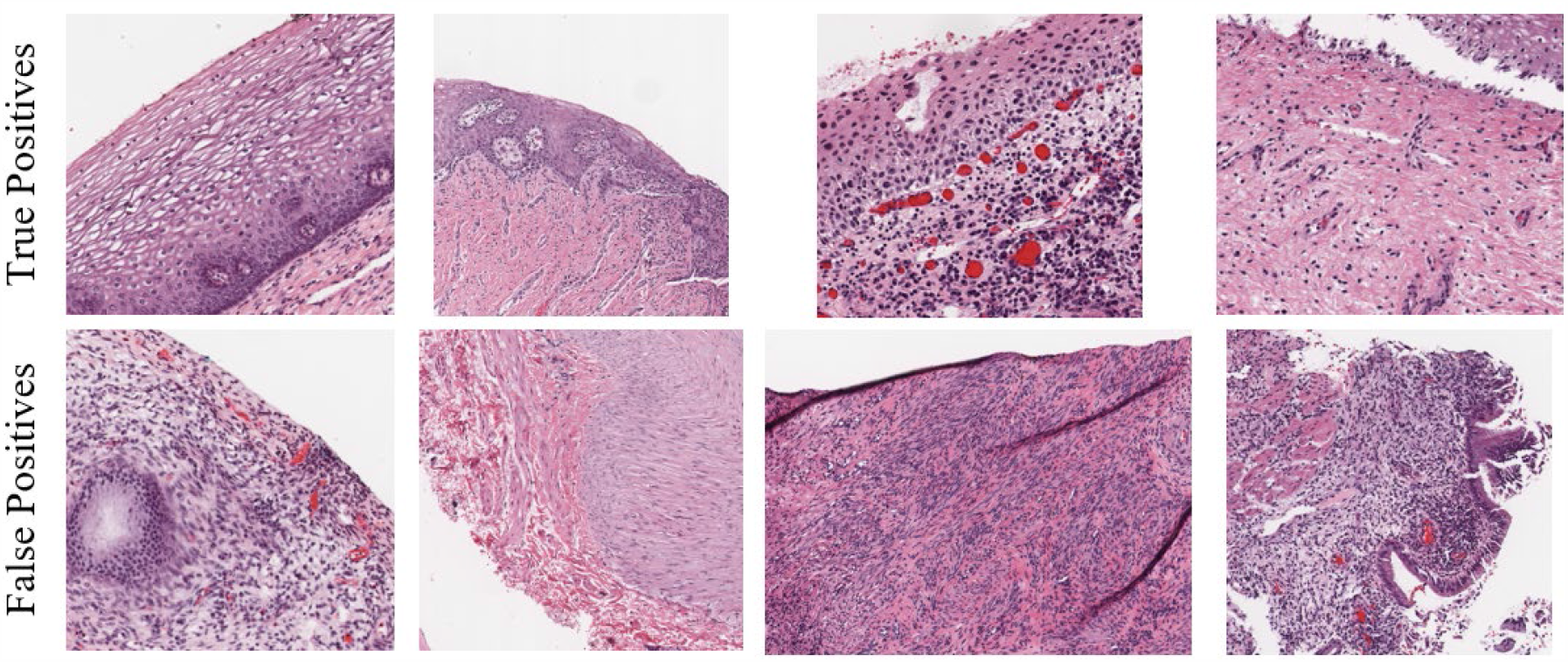
Examples of epithelium detection results. Correctly classified (top row) and misclassified (bottom row) epithelium ROIs.

### Performance of CIN Classification Model

We evaluate and compare the performance of the CIN classification model on the high-resolution epithelium images extracted through the proposed automated epithelium detection and segmentation process, and manually cropped and segmented images. We employ five scoring schemes [18] to analyze the classification results. They are exact class labels, CIN versus Normal, CIN3-CIN2 versus CIN1-Normal, CIN3 versus CIN2-CIN1-Normal, and off-by-one class.

The classification results from the DeepCIN classification model for OU15 and OU16 image sets are tabulated in Table 5 and Table 6, respectively. The results indicate that the DeepCIN model performed better on the OU16 dataset compared to the OU15 dataset. This may be due to the presence of relatively fewer artefacts during the preparation of the OU16 WSIs compared to OU15 WSIs. Table 7 shows the results of the combined dataset. We observe that the model has few misclassifications, usually off by one CIN grade. This can be observed by the off-by-one class results. This disagreement is also observed to happen among expert pathologists with interobserver variability. If we rank the scoring schemes based on the results, the off-by-one class is followed by CIN versus Normal (abnormal vs normal), which is helpful to distinguish the abnormal precancerous epithelium regions from the normal epithelium regions. These are followed by, CIN3-CIN2 versus CIN1-Normal, CIN3 versus CIN2-CIN1-Normal, and exact class labels.

**Table 5.** CIN classification results on OU15-set.

| Scoring Scheme | P | R | F1 | ACC | AUC | AP | MCC | $\kappa$ |
| --- | --- | --- | --- | --- | --- | --- | --- | --- |
| Exact class label | 83.1 | 83.8 | 82.8 | 83.8 | 94.4 | 86.8 | 70.35 | 70.1 |
| CIN vs Normal | 91.1 | 91.1 | 91.1 | 91.1 | 90.1 | 95.7 | 81.0 | 81.0 |
| CIN3-CIN2 vs<br>CIN1-Normal | 93.2 | 93.2 | 93.8 | 93.2 | 89.1 | 97.8 | 81.6 | 81.3 |
| CIN3 vs CIN2-CIN1-Normal | 93.6 | 94.2 | 92.8 | 94.2 | 63.7 | 95.5 | 46.1 | 39.4 |
| Off-by-one | - | - | - | 96.3 | - | - | - | - |

**Table 6.** CIN classification results on OU16-set.

| Scoring Scheme | P | R | F1 | ACC | AUC | AP | MCC | $\kappa$ |
| --- | --- | --- | --- | --- | --- | --- | --- | --- |
| Exact class label | 90.2 | 88.4 | 88.2 | 88.4 | 98.0 | 93.1 | 80.5 | 80.0 |
| CIN vs Normal | 97.3 | 97.3 | 97.3 | 97.3 | 97.2 | 99.7 | 94.4 | 94.4 |
| CIN3-CIN2 vs<br>CIN1-Normal | 95.7 | 95.6 | 95.5 | 95.5 | 94.0 | 99.1 | 90.3 | 90.0 |
| CIN3 vs CIN2-CIN1-Normal | 93.0 | 92.4 | 91.5 | 92.4 | 78.2 | 97.0 | 71.9 | 68.1 |
| Off-by-one | - | - | - | 98.2 | - | - | - | - |

**Table 7.** CIN classification results on the combined set.

| Scoring Scheme | P | R | F1 | ACC | AUC | AP | MCC | K |
| --- | --- | --- | --- | --- | --- | --- | --- | --- |
| Exact class label | 85.0 | 85.0 | 84.2 | 85.0 | 95.5 | 88.3 | 73.0 | 72.7 |
| CIN vs Normal | 92.6 | 92.6 | 92.6 | 92.6 | 92.0 | 96.9 | 84.3 | 84.3 |
| CIN3-CIN2 vs<br>CIN1-Normal | 93.8 | 93.8 | 93.7 | 93.8 | 90.5 | 98.3 | 84.1 | 83.9 |
| CIN3 vs CIN2-CIN1-Normal | 93.7 | 93.8 | 92.6 | 93.8 | 69.7 | 96.0 | 58.3 | 52.9 |
| Off-by-one | - | - | - | 96.7 | - | - | - | - |

The proposed toolbox is observed to face difficulty in correctly identifying the CIN 3 epithelium images. There is an off-by-one grade error with misclassification as CIN 2. This can be observed from the metric values of CIN3-CIN2 versus CIN1-Normal and CIN3 versus CIN2-CIN1-Normal scoring schemes in Table 7.

The performance of the proposed toolbox for automated cervical diagnosis is benchmarked against CIN classification results on the manually cropped and segmented epithelium images (Table 8). The manually extracted epithelium images were chosen carefully to capture and focus on the epithelium regions along with accurate annotations for epithelium masks. These images are close to the ideal conditions, and we compare them with the epithelium images from an automated realistic toolbox. We observed that the proposed toolbox has a closer performance to the benchmark results, and this indicates that the proposed prototype has the potential to be used in real-world clinical settings.

**Table 8.**
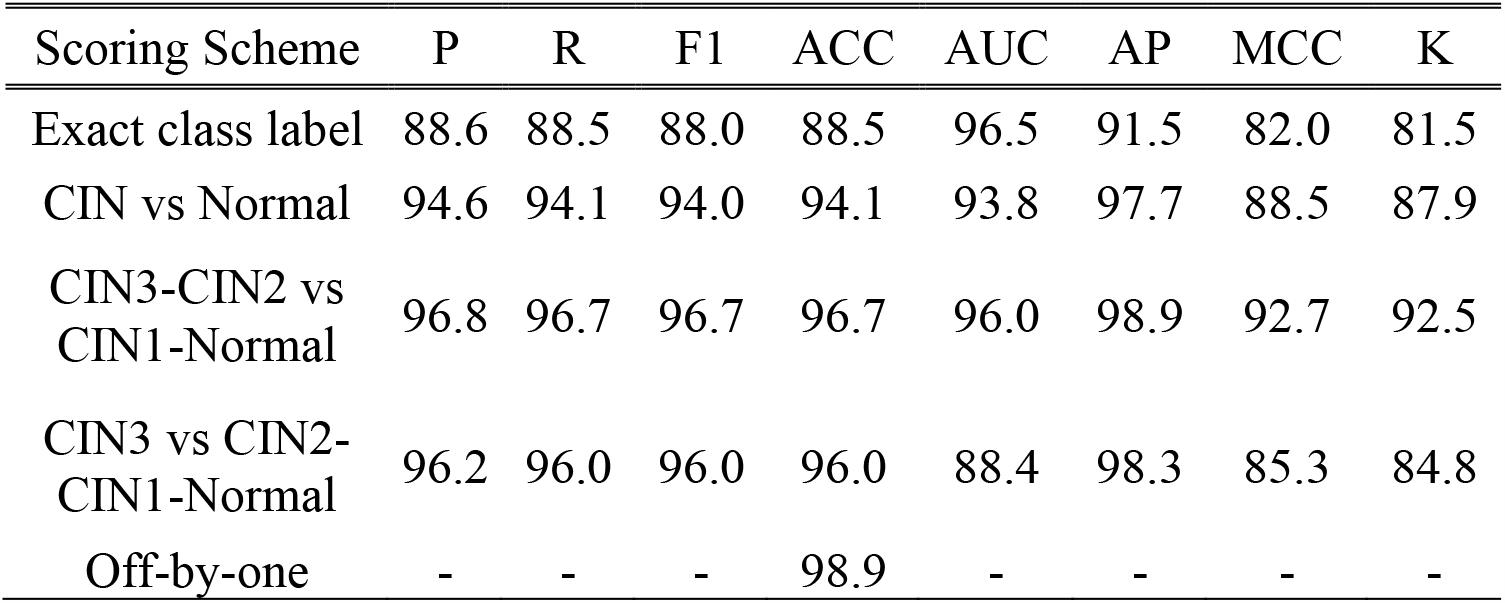
Benchmark CIN classification results [18].

| Scoring Scheme | P | R | F1 | ACC | AUC | AP | MCC | K |
| --- | --- | --- | --- | --- | --- | --- | --- | --- |
| Exact class label | 88.6 | 88.5 | 88.0 | 88.5 | 96.5 | 91.5 | 82.0 | 81.5 |
| CIN vs Normal | 94.6 | 94.1 | 94.0 | 94.1 | 93.8 | 97.7 | 88.5 | 87.9 |
| CIN3-CIN2 vs<br>CIN1-Normal | 96.8 | 96.7 | 96.7 | 96.7 | 96.0 | 98.9 | 92.7 | 92.5 |
| CIN3 vs CIN2-CIN1-Normal | 96.2 | 96.0 | 96.0 | 96.0 | 88.4 | 98.3 | 85.3 | 84.8 |
| Off-by-one | - | - | - | 98.9 | - | - | - | - |

## V. DISCUSSION

The cervical histopathology data suffers from three major limitations in the data collection and preparation. Firstly, unlike scenic images used in the public challenges, biomedical image data requires a lot of approvals to gather patient data and hence the amount of data is relatively very low. Secondly, gathering expert labelled data is always challenging since this requires skills to identify the regions and grade the cancer. There is always an inter-pathologist variation related to interpretation of the results. A study [22] has shown that there is an interobserver variability of 0.799 to 0.887 in terms of kappa score among four expert pathologists who have CIN grading experience of 8-30 years. Thirdly, the distribution of the data is always skewed/ imbalanced as shown in Table 2 and Table 3.

This paper is intended to compare the proposed automated digitized histology slide analysis for CIN classification of epithelium regions with the manually segmented epithelium regions. Fewer epithelium regions in WSIs were considered for evaluation due to limited availability of expert pathologist labeling. Future studies will explore the interpretation of WSI-level CIN classification for the complete end-to-end digitized slide analysis. The inclusion of techniques like graph theory for deeper understanding of spatial context and data fusion might help in further improving the CIN classification results. Stacked models can be created to handle the lack of consensus pathology, that is, the designed models should have the ability to interpret the disagreements among the pathologists’ ground truth labelling. The resolution of WSI scanners should be a concern too. There is variability across manufacturers which leads to issues with different image resolutions. The future work will be also be focused on designing models that can handle WSIs from various sources.

## VI. CONCLUSION

Our pipeline draws inspiration from the examination strategy of an expert pathologist, where he/she scrutinizes the growth of abnormal cells across small portions of the epithelium. This is realized by scanning the cervical histopathological WSI and extracting the epithelium ROIs present on the outer layer of the tissue sample. Since there are regions without epithelium, filtering the ROIs is crucial to retain only the epithelium ROIs and this is accomplished by the proposed epithelium detection network. With the help of our previous studies, we incorporated the EpithNet-64 model for segmenting the epithelium regions in the epithelium ROIs. Small vertical portions are extracted for a localized cell growth pattern analysis, which is performed by the DeepCIN model. The results sequences are fused with attentional observation to determine the final CIN grade for the epithelium ROI. Even the significance of the local regions was identified in this process of CIN classification. Furthermore, the CIN grade for the entire WSI can be generated by voting CIN classification results from the portions of epithelium ROIs.

We observed that our unique, novel approach for an automated CIN diagnosis from a WSI has achieved expert pathologist level accuracy. This clearly indicates the potential of our proposed pipeline as an assisting tool to an expert pathologist both in terms of quality of diagnosis and time. Due to the limitation of the data samples and expert annotated WSI-level labels, we tend to quote the toolbox as a prototype. If there is the availability of more data from various sources, the toolbox could be better generalized for use by everyone. The tool can be further improved by considering additional information of patients’ metadata and genetic codes.

## Data Availability

The data is used for research purposes in collboration with NIH and haven't been made available to the public.

## Acknowledgments

This research was supported in part by the Intramural Research Program of the National Institutes of Health (NIH); National Library of Medicine (NLM); and Lister Hill National Center for Biomedical Communications (LHNCBC). In addition, we gratefully acknowledge the medical expertise of Dr. P. Deepika Devada.

